# An outbreak investigation of a large dengue fever outbreak in Somaliland, 2023–2024

**DOI:** 10.1101/2025.06.13.25329553

**Authors:** H Asad, A Ali, A Hashi, I Hirsi, M Bahdoon, VJ Del Rio Vilas, OJ Brady, H Bower, F Haque, R Li, M Hergeye

## Abstract

Somaliland experienced a large dengue fever outbreak in 2023–2024, with 4,971 probable cases, 1,703 confirmed by rapid tests, and three deaths. Reported cases spanned five regions, with 74% in Marodijeh. We highlight the utility of the new laboratory surveillance and the need for mitigating dengue risks amid climatic fluctuations.

## Introduction

Dengue fever (DF), transmitted primarily by the *Aedes aegypti* mosquito, has significantly impacted Eastern Africa and the Middle East since re-emergence in late 20^th^ century: with three serotypes now circulating (DENV-1, DENV-2, DENV-3) (2–5). Somaliland is a self-declared independent region in Eastern Africa since 1991. Its first reported DF outbreak was in its capital city Hargeisa during 1985–87 (6). No further outbreak was reported (3) until October 2022, when the Somaliland Ministry of Health Development (MOHD) declared a DF outbreak (7) concurrent with outbreaks in neighbouring regions of Somalia (8,9). By February 2023, fever cases at public healthcare facilities in Somaliland declined and remained low (Appendix 1), until 14/09/2023 when a new cluster of probable dengue patients was identified at a hospital near Hargeisa. On confirmation by the National Public Health Laboratory (NPHL) using reverse transcriptase polymerase chain reaction (RT-PCR), MOHD declared a new DF outbreak on 03/10/2023 and initiated control measures including spraying for vector reduction, risk communication and community engagement, and training of healthcare personnel on clinical management. On 21/10/2023, MOHD requested assistance from UK Public Health Rapid Support Team to conduct a joint epidemiologic investigation.

## Methods

### Case definition

A “probable DF case” was defined as having fever and ≥2 of the following symptoms: nausea/vomiting, rash, aches and pains, tourniquet test positive, leukopenia, any warning sign (10), identified at a healthcare facility in Somaliland, with symptom onset between 01/09/2023 and 30/06/2024. A “confirmed case” was a probable case with positive result for immunoglobulin M (IgM) or NS1 antigen in a single serum sample using a rapid diagnostic test (RDT). Sample collection or reporting date was assumed to represent symptom onset date if unavailable.

### Case finding and data source

We collected morbidity data from NPHL and mortality data from MOHD. From September 2023, NPHL procured a limited supply of dengue RDTs (SD Bioline Dengue Ag /Ab (NS1 / IgM)) from the World Health Organization (WHO) and trained personnel in public-sector referral health centers and hospitals to test probable patients and report all tests to a newly established surveillance system with data on: date of sample collection, type and result of dengue test, patient’s gender, age, place of residence, presenting healthcare facility, presenting symptoms, and date of symptom onset. Only probable cases that underwent laboratory testing were included in the system. Data were received from seven of the 23 districts spanning five of Somaliland’s six regions (Figure 1); Sool Region did not report dengue testing due to unstable situation. MOHD collated the number of dengue-related deaths retrospectively.

### Data analysis

We report the attack rate (number of probable or confirmed cases/population based on the 2023 MOHD Population Estimation Survey) and the case fatality rate (CFR) (number of dengue-related deaths/number of probable cases).

### Ethics

Ethical approval was not required as analyses were part of monitoring and response to an ongoing outbreak.

## Results

Between 01/09/2023 and 30/06/2024, Somaliland NPHL documented 4971 probable DF cases, with 1703 (34.2%) confirmed by RDT (**Table**). The outbreak predominantly affected younger adults and children (median age of all cases: 23 years; interquartile range: 10–35). Overall DF attack rate was 109 cases per 100,000 population over the 10-month period, with 37.2 confirmed cases per 100,000. MOHD reported three DF-related deaths (CFR 0.06%).

The outbreak peaked in mid-November 2023 with 436 new cases per week, predominantly from the capital region of Marodijeh (**Figure A-C**). Cases dropped to fewer than 100 weekly cases in early 2024. Between January and May 2024, the northeastern regions of Sahil and Sanag were most affected.

Marodijeh experienced a second surge in June 2024. Throughout the outbreak, Togdheer and Awdal region reported only intermittent cases.

Patient characteristics varied by time and region (**Appendix Table 1**). Marodijeh saw older cases in the 2024 wave compared to the 2023 wave (median age 25 vs 18 years, Mann-Whitney test *p* <.001). Other regions reported median ages ranging from 25 to 32 years. Gender distributions were relatively balanced (48.5% female). Similar patterns were observed among confirmed cases (**Appendix Table 2**).

RDT testing began in September 2023 in Marodijeh, expanding to all regions except Sool by November 2023. Overall, one third of probable cases tested positive for dengue (**Table, Figure D-E**). Older patients were more likely to test positive (chi-squared trend test: *p* =.004). Test positivity exceeded 40% during the first two months of the outbreak and as cases resurged in May–June 2024. Frequent testing gaps were observed in Awdal, Sahil, Sanag, and Togdheer regions (**Appendix Figure 2**), coinciding with reported shortages of RDT kits.

## Discussion

This investigation describes extensive and recurring dengue transmission in Somaliland from September 2023 to June 2024 with 4,971 probable cases and three reported deaths. Concurrent transmission was reported in neighboring Ethiopia (11,12) and Yemen (4). While clinical features of the second wave of this outbreak have previously been described (13), here we show that transmission was sustained across multiple waves for almost a year and spread across multiple regions. The presence of cases across all age groups, matching the population’s age structure, and the low case fatality rate (0.06%) suggest that most cases represent primary dengue infections. As subsequent infections with different dengue serotypes are more likely to have serious clinical manifestations, this leaves Somaliland vulnerable to potentially more severe future outbreaks.

Recent extreme climatic conditions in Eastern Africa may have contributed to changes in dengue risk, as high rainfall or droughts that lead to standing water in open containers increase *Aedes Aegypti* egg-laying habitat (14,15). Historically, Somaliland observes two rainy seasons: March–June (*gu*) and October–November (*deyr*) (16). The 2023 *deyr* rains were exceptionally heavy (17), coinciding with the outbreak’s onset. Rainfall deficits in the 2024 *gu* season (18) coincided with a new peak in dengue cases in June. These fluctuations underscore the urgent need for integrated surveillance combining entomological and meteorological data to better understand dengue transmission dynamics in the region. Drought and flood response planning in Somaliland must specifically address potential increases in dengue risk and identify strategies for mitigation.

The investigation has several limitations. Reliance of NPHL’s surveillance on laboratory-tested probable cases led to underreporting of milder cases, cases from areas with limited healthcare access (one third of population lives in nomadic settlements, **Appendix Table 3**), and cases presenting to primary-care facilities, which had very limited or no access to case detection training nor RDT kits. Changing availability of RDT affected clinical criteria for testing; no data were available to estimate testing fraction. Syndromic surveillance will complement RDT testing and mitigate this bias in resource-constrained settings. Additionally, NPHL did not have access to data from private facilities; the 2020 Somaliland Health and Demographic Survey reported residents were 50% more likely to visit private facilities than public ones when sick (19). Furthermore, the retrospective reporting of dengue-related deaths and lack of information on patient outcome in laboratory surveillance may have resulted in underestimation of the true CFR; a CFR of 0.06% is low compared to those reported by some other regions (0.01–1%) (2). Lastly, the absence of vector and serotype surveillance and reporting of tests for other pathogens of probable cases who were RDT negative prevented more comprehensive analyses of outbreak dynamics and potential changes in severity.

Despite these limitations, the investigation demonstrates the utility of the new laboratory surveillance system in Somaliland during its first year of operation. Strengthening an integrated DF surveillance systems will not only preserve institutional memory but also provide insights into how socioeconomic and climatic factors amply dengue risks. Given the large nomadic population, understanding the spatiotemporal patterns of disease burden is crucial for informing more tailored interventions to mitigate dengue transmission.

## Data Availability

All data produced in the present study are available upon reasonable request to the authors

## Acknowledgments

We thank healthcare facilities in Somaliland for caring for dengue patients and reporting data to the National Public Health Laboratory surveillance. HA thanks partners in the Somaliland MoHD for their collaboration and operational colleagues in the UK Public Health Rapid Support Team and Foreign, Commonwealth & Development Office for supporting her deployment in December 2023–March 2024. The UK Public Health Rapid Support Team is funded by UK Aid from the Department of Health and Social Care and is jointly run by the UK Health Security Agency and the London School of Hygiene and Tropical Medicine. The views expressed in this publication are those of the author(s) and not necessarily those of the Department of Health and Social Care.

---

**Figure:**
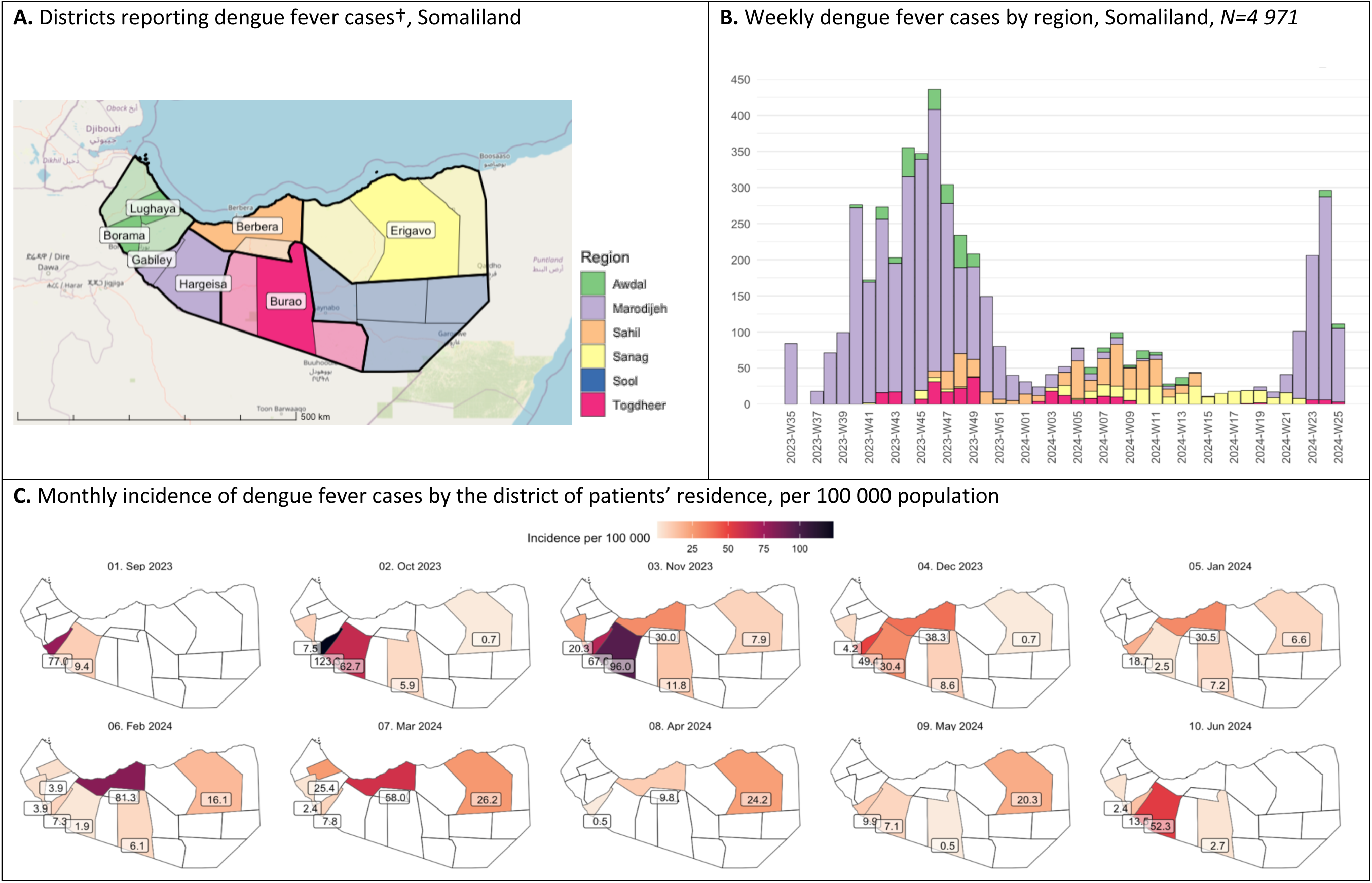
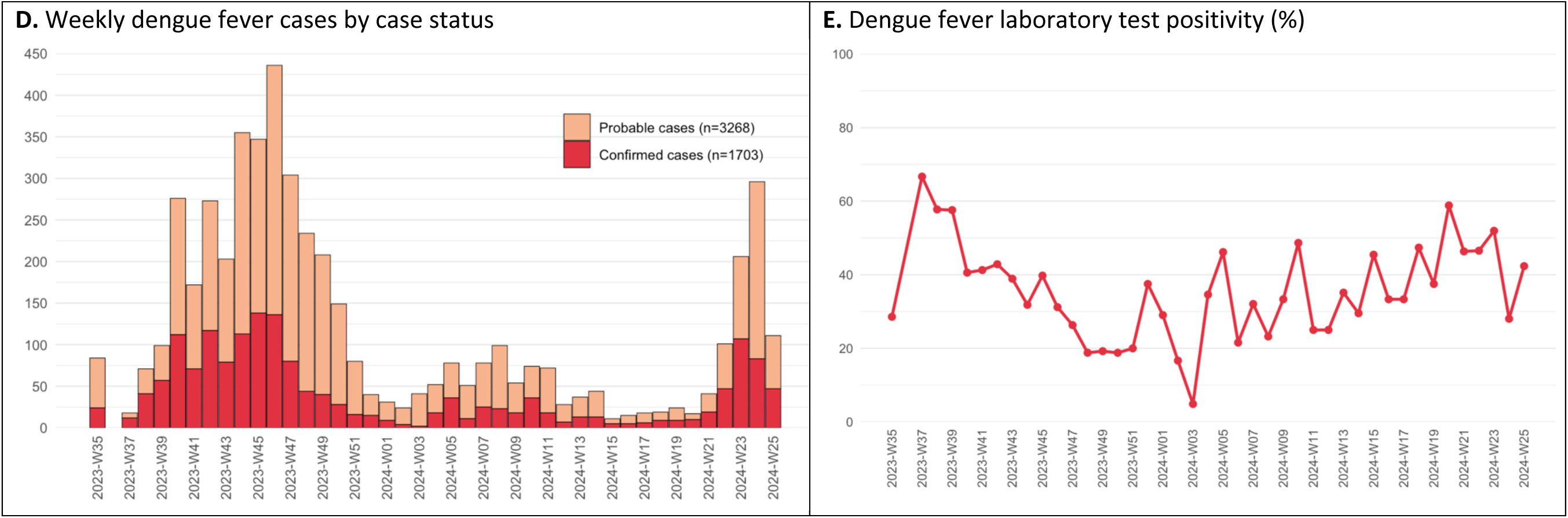
Distribution of dengue fever cases, Somaliland, 1 September 2023 *–* 30 June 2024**All probable and confirmed dengue fever cases. Dates shown on the x-axes are the first day of the epidemiologic week of symptom onset; if the date of onset is missing, the date of sample collection is used; if both dates are missing, the date of report is used. †Somaliland district borders are shown and districts colored based on Somaliland administrative regions. Districts reporting dengue fever cases are darker with district name labeled: Borama and Lughaya in Awdal Region; Gabiley and Hargeisa in Marodijeh; Berbera in Sahil; Erigavo in Sanag; and Burao in Togdheer. All patients were tested by a healthcare facility in the district of their residence, with one exception: some patients residing in Berbera district (Sahil region) were tested in Gabiley (Marodijeh region) between November and December 2023. Throughout the figures, cases were represented by patients’ district of residence.

**Appendix Figure 1:**
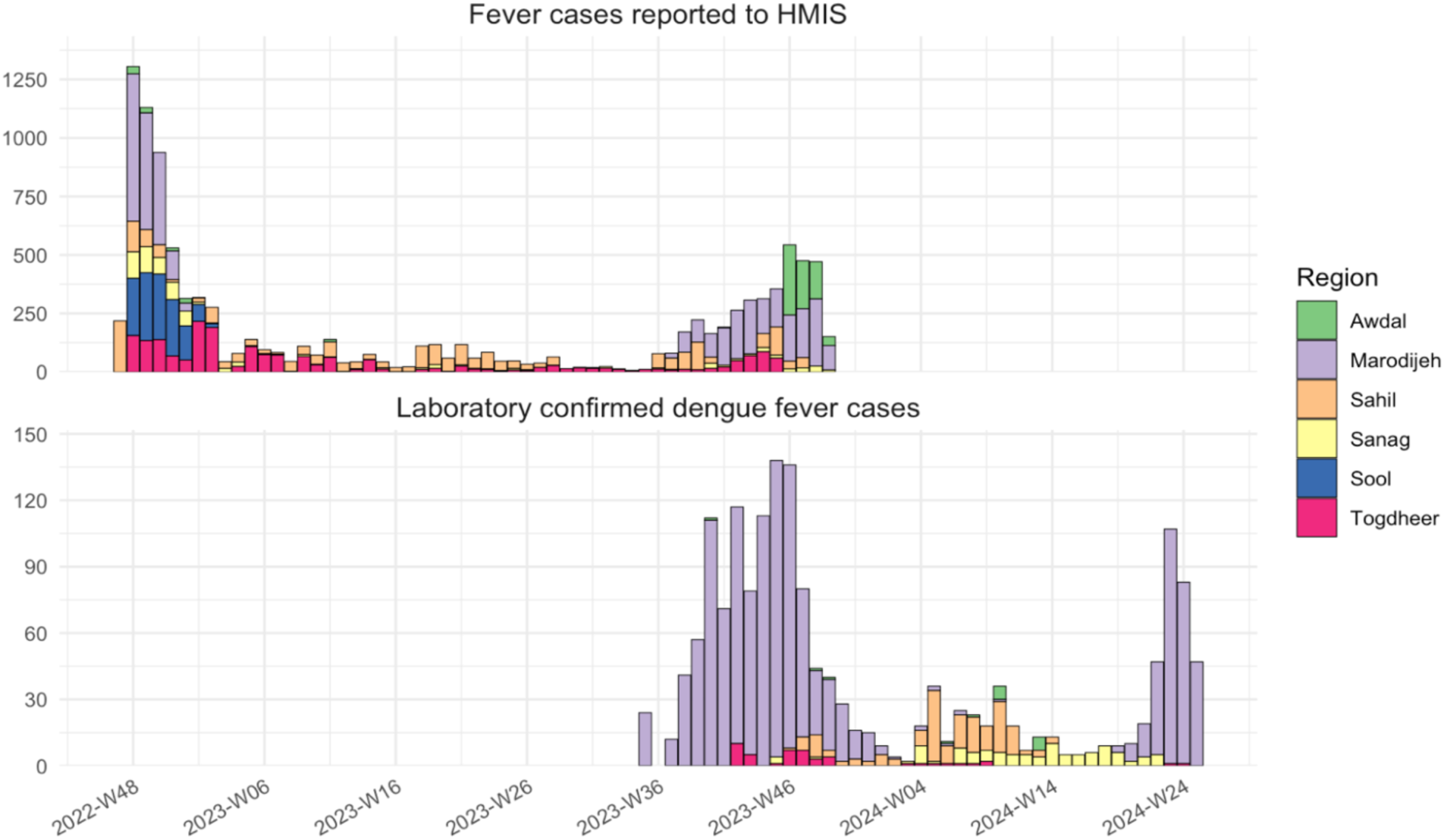
Weekly number of fever cases reported to HMIS*, Somaliland, 26 November 2022 – 6 December 2023 **\*** HMIS: Somaliland’s Health Management Information System. A daily count of patients presenting to public healthcare facilities with fever was reported to HMIS. The weeks shown are epidemiological week of report to HMIS. Since the start of the second outbreak, HMIS reported 3787 fever cases between 01/09/2023 and 06/12/2023. For laboratory confirmed cases, this is a reproduction of Figure B in main text to illustrate the overlap in the two surveillance systems. The weeks shown are epidemiological weeks of symptom onset; if the date of onset is missing, the date of sample collection is used; if both dates are missing, the date of report is used.

**Appendix Figure 2:**
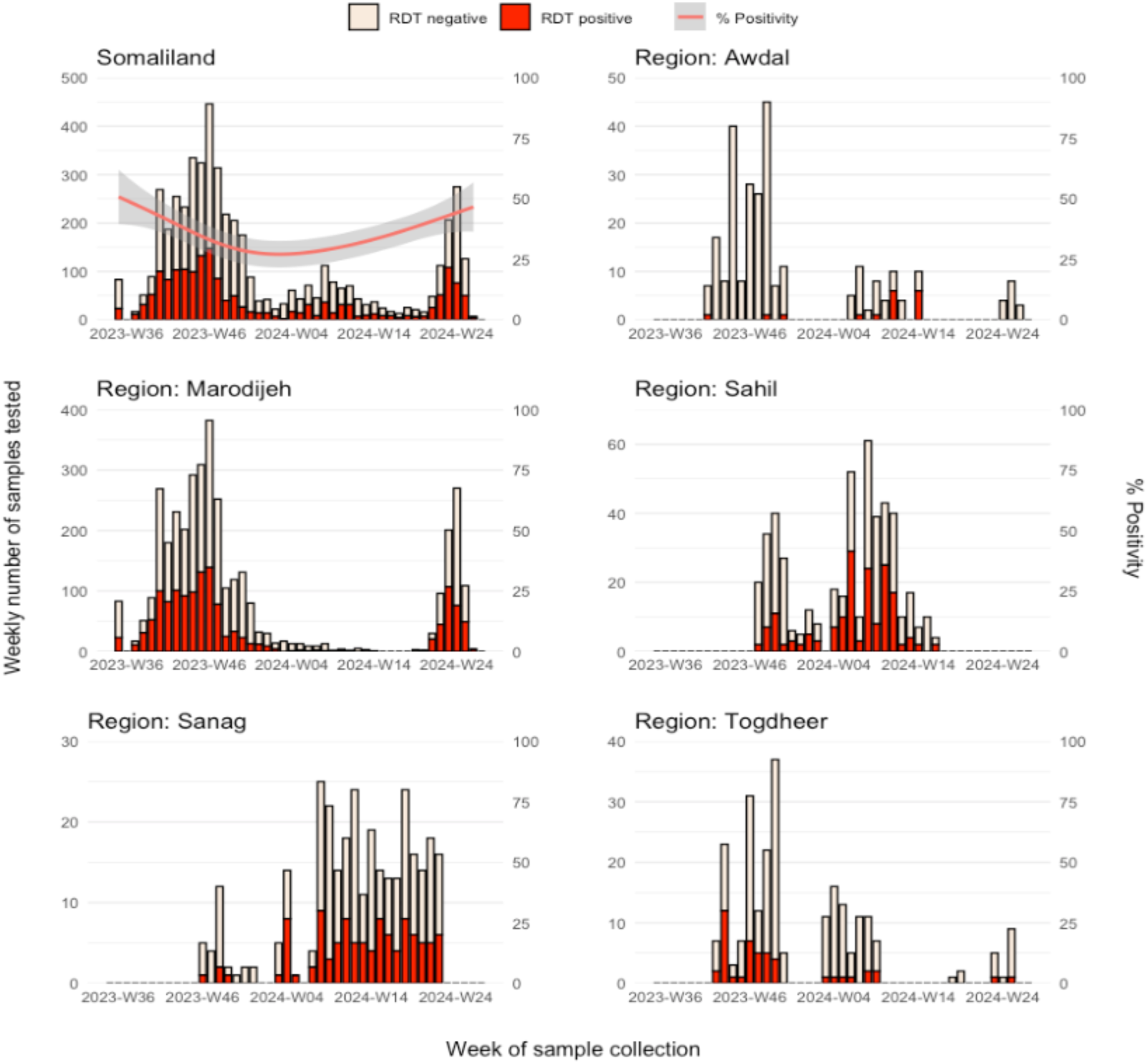
Trends in volume and positivity* of laboratory samples tested for dengue using rapid diagnostic tests, by region of patient residence†, Somaliland, 1 September 2023 – 30 June 2024‡ *Test positivity trend is fitted using a generalised additive model using cubic regression splines employing restricted maximum likelihood for optimal smoothing parameter selection, and is shown only for Somaliland due to small sample sizes and testing gaps for region-specific trends. †There are six regions in Somaliland; Sool did not report any dengue RDT testing, nor any healthcare facility tested patients residing with residence in Sool. In Awdal, two districts reported dengue RDTs (Borama, Lughaya); in Marodijeh, two districts (Gabiley, Hargeisa); in Sahil, one district (Berbera); in Sanag, one district (Erigavo), in Togdheer, one district (Burao). All patients were tested by a healthcare facility in the district of their residence, with one exception: some patients residing in Berbera district (Sahil region) were tested in Gabiley (Marodijeh region) between November and December 2023. ‡If date of sample collection is missing, date of report is used.

**Table:**
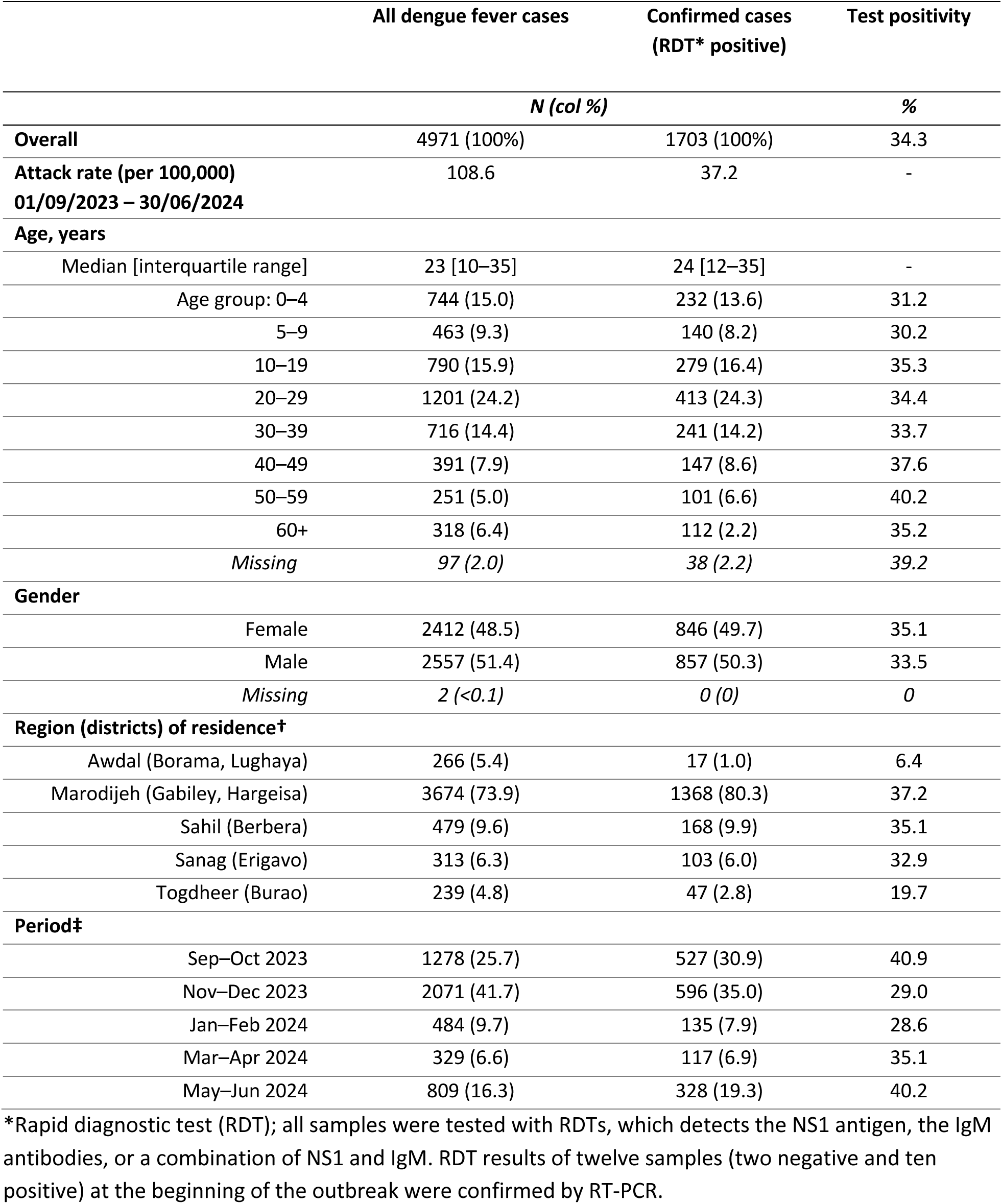
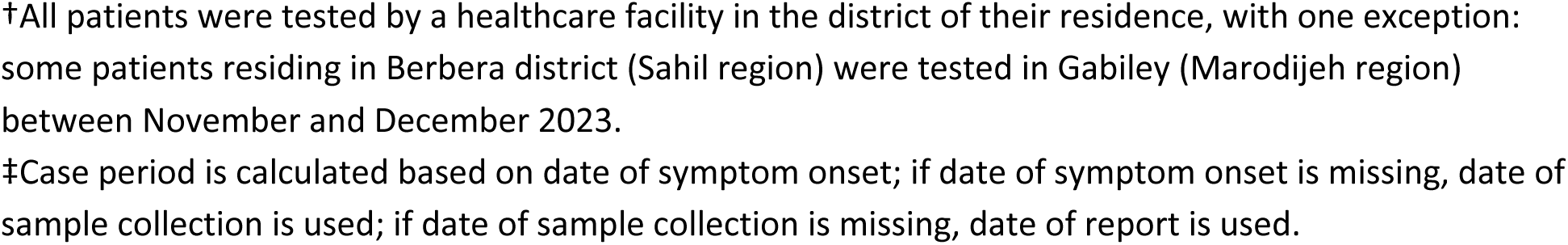
Dengue fever patient characteristics by rapid diagnostic test* positivity, Somaliland,1 September 2023–30 June 2024.

**Appendix Table 1:**
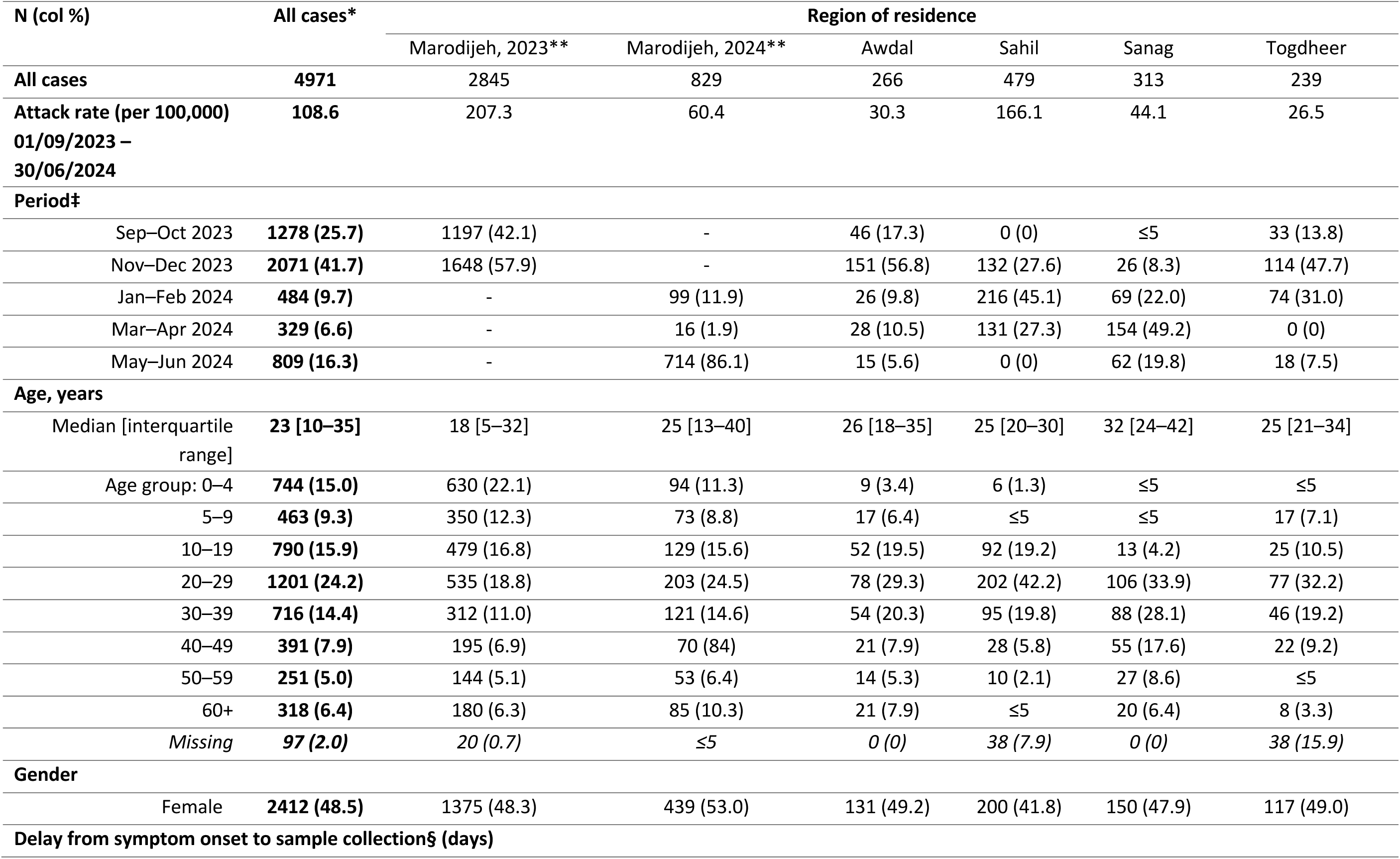
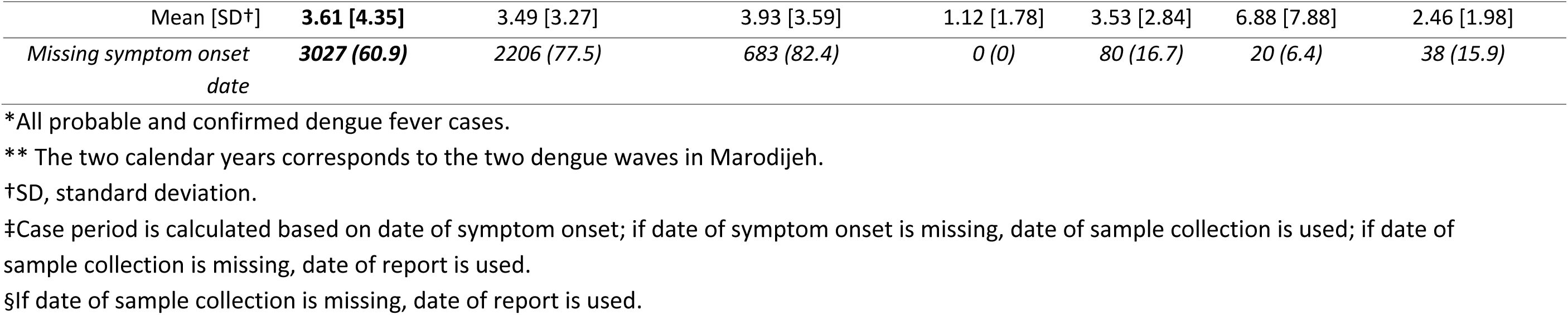
Characteristics of dengue fever cases* by region of residence, Somaliland, 1 September 2023 *–* 30 June 2024.

**Appendix Table 2:**
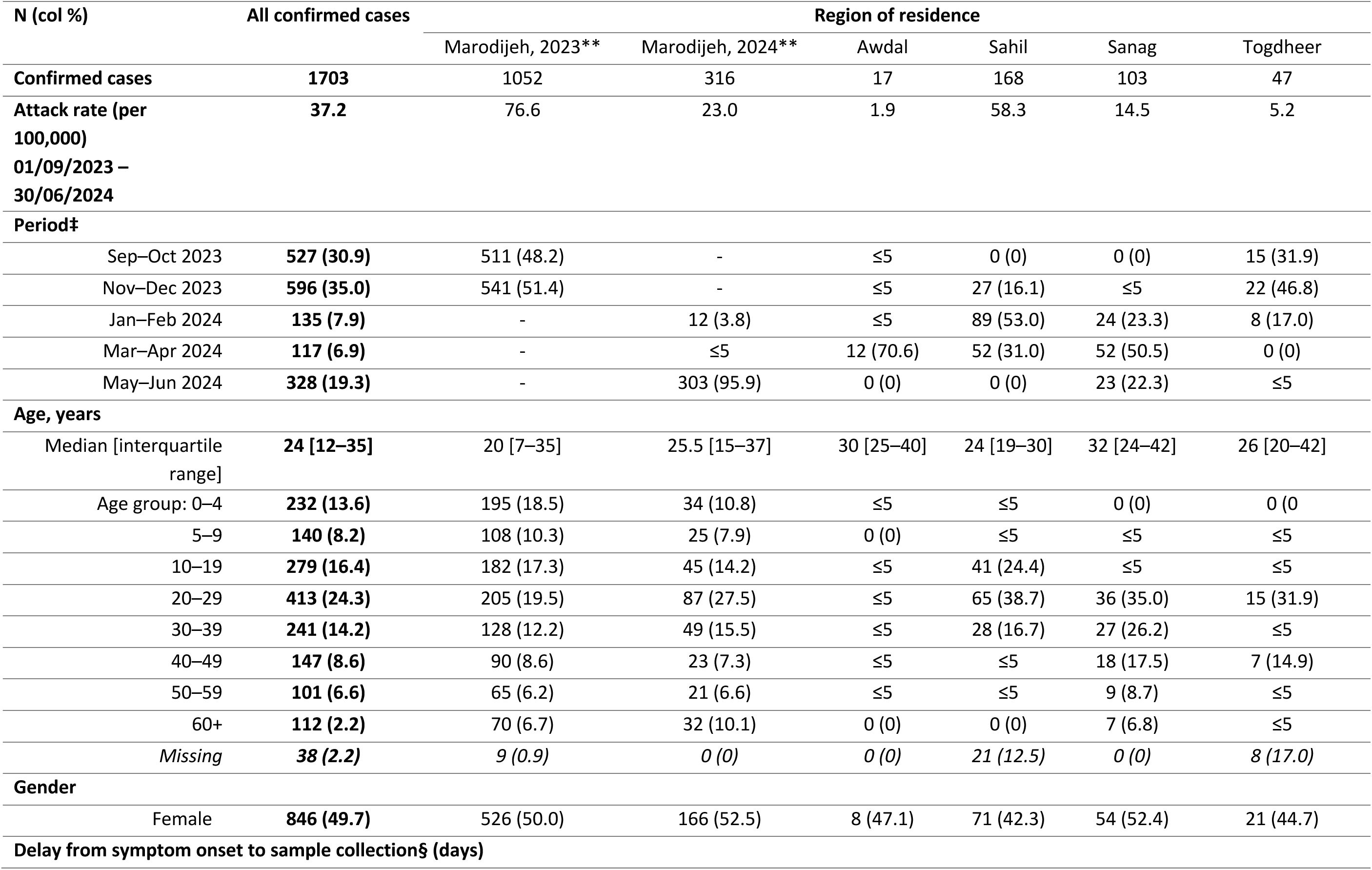
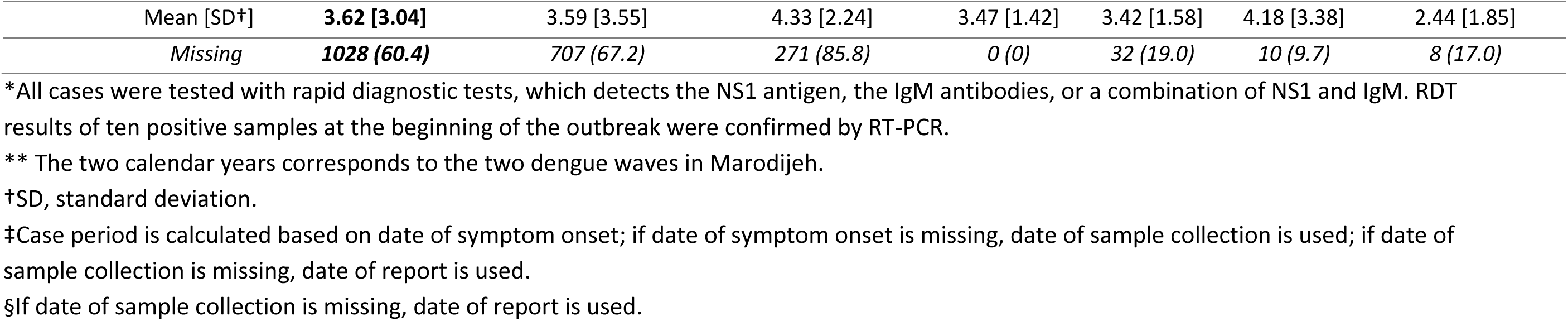
Characteristics of confirmed dengue fever cases*, Somaliland regions, 1 September 2023 *–* 30 June 2024.

**Appendix Table 3:** Somaliland Population by Type of Residence, 2016.

| Region<br>(Somaliland) | Region<br>(Somalia) | Population by type of residence*<br>(%) |  |  |  |
| --- | --- | --- | --- | --- | --- |
|  |  | Nomadic | IDP camps | Urban | Rural |
| Awdal | Awdal | 34.7 | 1.2 | 42.8 | 21.4 |
| Marodijeh + Sahil | W. Galbeed | 20.6 | 3.6 | 64.6 | 11.2 |
| Togdheer | Togdheer | 21.4 | 3.6 | 67.1 | 8.0 |
| Sool | Sool | 57.3 | 1.5 | 37.0 | 4.3 |
| Sanag | Sanaag | 64.8 | 0.2 | 29.4 | 5.7 |
| **Total Somaliland\*\*** |  | **33.7** | **2.4** | **52.9** | **11.0** |

IDP, internally displaced population.
\* Estimates based on *UNFPA (2016). Population Composition and Demographic Characteristics of the Somali People, Volume 2, Figure 19, page 22.*
\*\*Calculated based on the region-specific proportion of population by type of residence (*Figure 19, page 22*) and the population size as a proportion of total Somalia population (*Table 2, page 20*).

## Notes

### Competing Interest Statement

The authors have declared no competing interest.

### Author Declarations

Ethics committees of Londons School of Hygiene and Tropical Medicine, UK Health Security Agency, and Somaliland Ministry of Health Development waived ethical approval for this work as analyses were part of monitoring and response to an ongoing outbreak.

